# Development of a multiplex real-time PCR assay for BCG and validation in a clinical laboratory

**DOI:** 10.1101/2021.06.04.21258161

**Authors:** Shannon C Duffy, Manigandan Venkatesan, Shubhada Chothe, Indira Poojary, Valsan Philip Verghese, Vivek Kapur, Marcel A Behr, Joy S Michael

## Abstract

*Mycobacterium bovis* bacille Calmette-Guérin (BCG) is a live attenuated vaccine which can result in local or disseminated infection, most commonly in immunocompromised individuals. Differentiation of BCG from other members of the *Mycobacterium tuberculosis* complex (MTBC) is required to diagnose BCG disease, which requires specific management. Current methods for BCG diagnosis are based on mycobacterial culture and conventional PCR; the former is time-consuming and the latter often unavailable. Further, there are reports that certain BCG strains may be associated with a higher rate of adverse events. This study describes the development of a two-step multiplex real-time PCR assay which uses single nucleotide polymorphisms to detect BCG and identify early or late BCG strains. The assay has a limit of detection of 1 pg BCG boiled lysate DNA and was shown to detect BCG in both pure cultures and experimentally infected tissue. Performance was assessed on 19 suspected BCG clinical isolates at Christian Medical College in Vellore, India taken from January 2018 to August 2020. Of these 19 isolates, 10 were identified as BCG (6 early and 4 late strains) and 9 were identified as other MTBC members. Taken together, the results demonstrate the ability of this assay to identify and characterize BCG disease from cultures and infected tissue. The capacity to identify BCG may improve patient management and the ability to discriminate between BCG strains may enable BCG vaccine pharmacovigilance.

## Introduction

*Mycobacterium bovis* bacille Calmette-Guérin (BCG) is a live attenuated vaccine which is widely administered for the protection against tuberculosis^1^. BCG vaccination can be associated with local adverse events such as injection site abscesses or lymphadenitis. It may also lead to systemic infection causing osteomyelitis or disseminated BCG^2^. Disseminated disease is rare and often occurs in young children with primary immunodeficiencies (PID) such as severe combined immunodeficiency (SCID), chronic granulomatous disease (CGD), and mendelian susceptibility to mycobacterial disease (MSMD), or those with HIV^3, 4^. While BCG vaccination is contraindicated in people known to have these conditions, for programmatic reasons, BCG is given to neonates, such that these immunodeficiencies are typically only identified until after vaccination has occurred^1^.

To diagnose BCG disease, laboratory differentiation of mycobacterial isolates is essential, as BCG has similar growth kinetics and morphology to *M. tuberculosis* strains. Current methods of diagnosis of BCG often rely on detection of regions of difference (RD) by PCR^5-10^. Recent evidence has suggested that the use of deletions for differentiation of *Mycobacterium tuberculosis* complex (MTBC) subspecies may be unreliable^11^ as several regions of the genome are hotspots of independent deletion events^12-14^. Furthermore, conventional gel-based PCR is not a pragmatic option in many clinical labs and can only target one region of the genome at a time.

Given the increased data on *M. tuberculosis* single nucleotide polymorphisms (SNPs) and the availability of real-time PCR reagents that enable multiplexed analyses, we developed a two-step real-time PCR assay to rapidly identify BCG from bacterial culture or directly from clinical samples. The two steps were designed to: 1) identify BCG from other members of the MTBC and 2) differentiate between early and late strains of BCG, something of interest in India where two different BCG strains (BCG Russia and BCG Danish) with potentially different rates of adverse events are in use. This assay was validated on 19 clinical isolates previously collected at Christian Medical College in Vellore, India and our results suggest that the use of this assay may better inform patient management and enable BCG vaccine pharmacovigilance.

## Materials and Methods

### Assay Design

A two-step real-time PCR protocol was designed to identify BCG isolates from clinical samples. Step one identifies BCG from other MTBC subspecies. It is a three-probe multiplex real-time PCR assay which detects the MTBC insertion element IS1081, the BCG-specific SNP *kdpD* c247t, and the pyrazinamide resistance SNP *pncA* c169g^15, 16^. Step two differentiates BCG isolates as early or late strains. It is a two-probe real-time PCR assay which detects the BCG early strain SNP in *crp* c140t and the BCG late strain SNP in *mmaA3* g293a^16^. The probes and primers were designed using Primer Express v. 3.0 (Applied Biosystems, Foster City, CA, USA). All primer and probe sequences are listed in table 1. The IS1081, kdpD, crp, and mmaA3 probes are TaqMan ® MGB probes (Applied Biosystems, Foster City, CA, USA) and the pncA probe is a PrimeTime® BHQ probe (Integrated DNA technologies, Coralville, IA, USA). All primers were purchased from Invitrogen (Carlsbad, CA, USA).

**Table 1:**
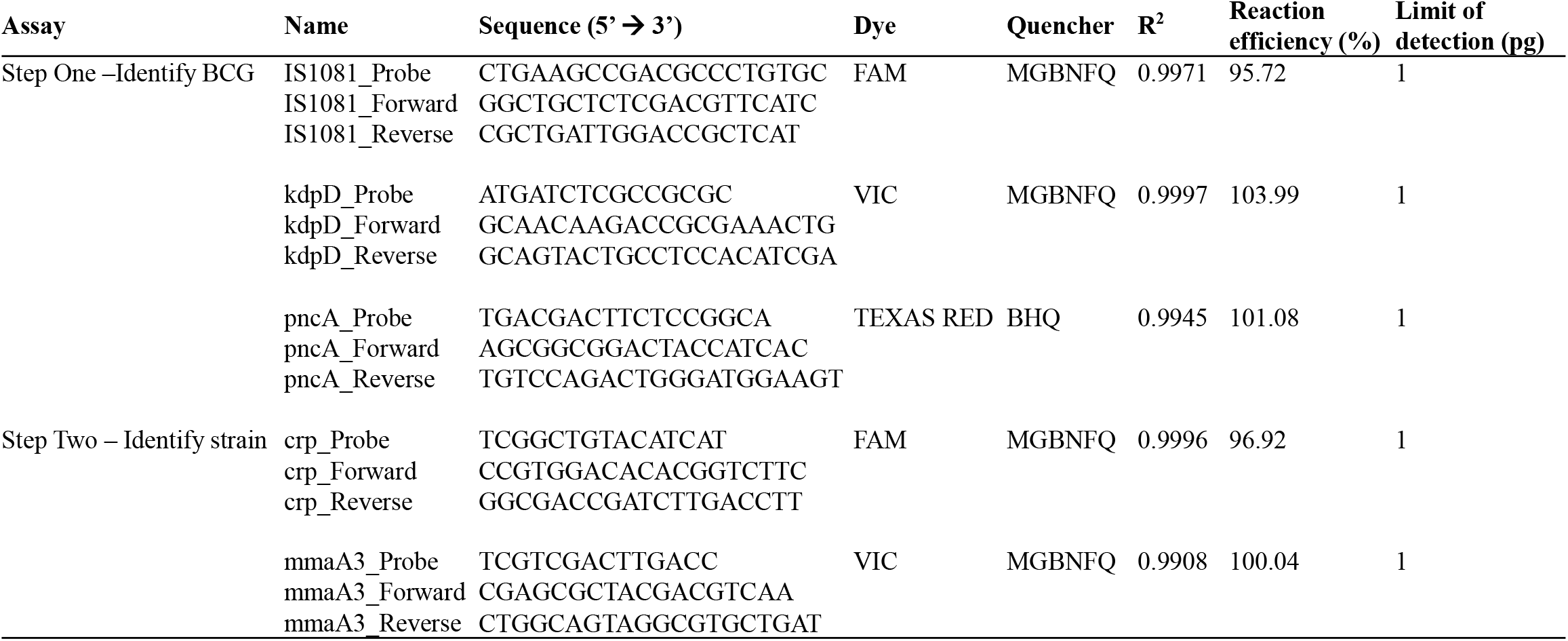
Sequences, reaction efficiencies, and limits of detection of probes and primers

### Real-time PCR

All real-time PCR reactions were performed in a 20 μl reaction containing 1 μl of DNA. Step one reaction mixtures were prepared with 10 μl TaqMan multiplex master mix (Applied Biosystems, Foster City, CA, USA), 100nM IS1081 forward and reverse primers, 300 nM forward and reverse kdpD primers, 900 nM pncA forward and reverse primers, 125 nM IS1081 probe, 125 nM kdpD probe, and 500 nM pncA probe. Step two real-time PCR reaction mixtures were prepared using 10 μl TaqMan multiplex master mix, 300 nM of crp and mmaA3 forward and reverse primers, and 250 nM crp and mmaA3 probes, and 1 μl dimethylsulfoxide (Sigma-Aldrich, St. Louis, MO, USA). Real-time PCR reactions were performed with a CFX96 touch real-time PCR detection system (Bio-Rad, Hercules, CA, USA) under the following thermocycling conditions: one cycle at 95°C for 10 minutes followed by 40 cycles of 95°C for 15 seconds and 63°C for 1 minute. A summary of results interpretation is found in figure S1.

### Specificity

The specificity of the two-step assay was evaluated with the following bacterial species: *M. tuberculosis, M. bovis, M. orygis* (2), *M. africanum, M. caprae* (2), *M. microti, M. avium* subsp. *paratuberculosis* (4), *M. avium* subsp. *avium* (3), *M. avium* subsp. *hominissuis, M. kansasii, M. abscessus, M. smegmatis*, BCG Russia, BCG Moreau, BCG Sweden, BCG Japan, BCG Danish, BCG Pasteur, BCG Prague, and BCG Glaxo. If more than one isolate of each mycobacterium was tested, the number of isolates is indicated in parentheses. Cultures of each bacterial species were grown in 7H9 Middlebrook media (Becton Dickinson, Franklin Lakes, NJ, USA), ADC (Becton Dickinson, Franklin Lakes, NJ, USA), 0.2% glycerol (Sigma-Aldrich, St. Louis, MO, USA), 0.05% Tween-80 (Sigma-Aldrich, St. Louis, MO, USA) to approximately an OD_600_ of 0.5-0.8. Media was supplemented with 1μg/mL ferric mycobactin J (Allied Monitor, Fayette, MO, USA) for *M. avium* subsp. *paratuberculosis* cultures. DNA was prepared from 1 mL culture by boiling. Boiled lysate DNA was used in this study to demonstrate assay robustness and evaluate the assay using a DNA preparation that would be a pragmatic and convenient option for many labs. Prepared DNA was stored at -20°C until ready for testing by real-time PCR.

### Reaction efficiencies, limits of detection, reproducibility, and robustness

The reaction efficiencies of all probes were determined by plotting the cycle threshold (Ct) values obtained from a standard curve of 10-fold dilutions of DNA isolated from BCG Russia or BCG Danish. The reaction efficiency was calculated using the formula *E* = (10^−1/*m*^ – 1) x 100% where *m* is the slope of the line. The limit of detection of all probes was determined by the minimum amount of DNA to yield a positive amplification result from the above standard curves. The inter- and intra-assay reproducibility was evaluated by comparing the Ct values of step one and step two assays completed on 3 separate plates with 5 technical replicates per plate. The ability of the assay to function in the presence of excess non-specific DNA was tested by performing real-time PCR with 1 ng of target DNA (BCG Russia or BCG Danish) and 10 ng of *M. abscessus* DNA.

### Mouse infection

The ability of the assay to identify BCG directly from tissue was determined using infected mice. Three C57BL/6 mice (Jackson Laboratories, Bar Harbor, MA, USA) were intravenously infected with 2.3 × 10^8^ CFU BCG Russia. After 3 weeks, mice were sacrificed, and liver and spleen samples were collected. Organs were homogenized and infection was confirmed by serial dilution and plating on Middlebrook 7H10 (Becton Dickinson, Franklin Lakes, NJ, USA), OADC (Becton Dickinson, Franklin Lakes, NJ, USA), 0.5% glycerol (Sigma-Aldrich, St. Louis, MO, USA) and PANTA (Becton Dickinson, Franklin Lakes, NJ, USA). DNA was extracted from organ homogenates as previously described^17^. DNA was then quantified and analyzed by the two-step real-time PCR assay.

### Clinical samples

The assay was validated using 19 clinical isolates previously collected at Christian Medical College, Vellore, India from January 2018 to August 2020. This study obtained institutional review board approval (approval number 11725, dated Dec 19, 2018) and ethical clearance from Christian Medical College. An isolate is defined as a pure microbe cultured from an individual patient. A sample is defined as the patient specimen provided for culture. Isolates were collected from patients presenting with suspected tuberculosis who were 2 years of age or younger or by request of the clinician due to suspected BCG disease. Clinicians suspected BCG involvement with a BCG site abscess or axillary lymphadenitis, or when disseminated MTBC was isolated in an infant with an immunodeficiency disorder such as SCID or MSMD. DNA was extracted from positive mycobacteria growth indicator tube (MGIT) or Lowenstein-Jensen (LJ) cultures as available. In one sample, culture was not available, and DNA was extracted directly from a pus swab. DNA was prepared by boiling or using the QIAamp DNA mini kit (Qiagen, Hilden, Germany). All DNA was analyzed by the step one real-time PCR assay. If an isolate was identified as BCG in step one, it was analyzed by step two.

## Results

### Specificity, reaction efficiency, and limit of detection

The specificity of the assay was tested on DNA from boiled lysate preparations of 27 isolates from 20 different mycobacterial species, including 8 BCG strains. The results for the step one and step two assays for all DNA correlated 100% as expected. The expected step one amplification curves of selected mycobacterial species are shown in figure 1. BCG was positive for all 3 probes (IS1081, kdpD, and pncA), *M. bovis* was positive for IS1081 and pncA, *M. tuberculosis* was positive for IS1081 only, and *M. abscessus* was negative for all 3 probes. The expected step two amplification curves of a BCG early and late strain are shown in figure 2. BCG Russia, an early strain, was positive for crp and BCG Danish, a late strain, was positive for mmaA3. The reaction efficiencies of probes were calculated by plotting the Ct values from a serial dilution of BCG Russia boiled lysate DNA for step one (Fig. S2) and both BCG Russia and BCG Danish boiled lysate DNA for step two (Fig. S3). The reaction efficiencies for all probes ranged between 95.72 – 103.99%, which are within the range of desired values of 90-110% (Table 1). The limit of detection of step one was 1 pg of BCG Russia boiled lysate DNA. For step two, the limit of detection was also 1 pg whether using BCG Russia or BCG Danish boiled lysate DNA (Table 1).

**Figure 1:**
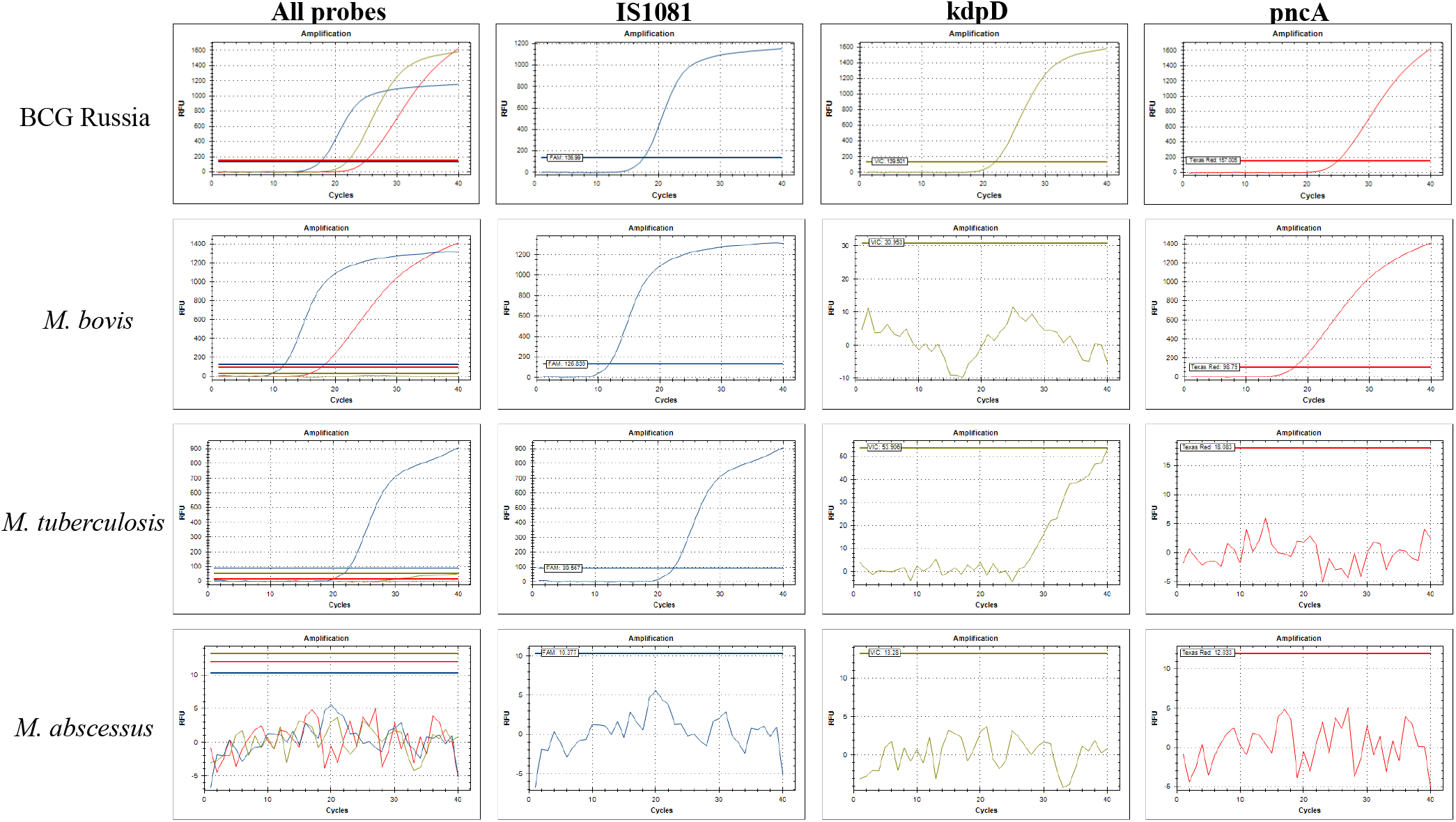
Step one assay specificity An example amplification curve for several mycobacteria with different probe amplification patterns: an MTBC subspecies *M. tuberculosis*, pyrazinamide-resistant *M. bovis*, a BCG strain BCG Russia, and a non-tuberculous mycobacterium *M. abscessus*.

**Figure 2:**
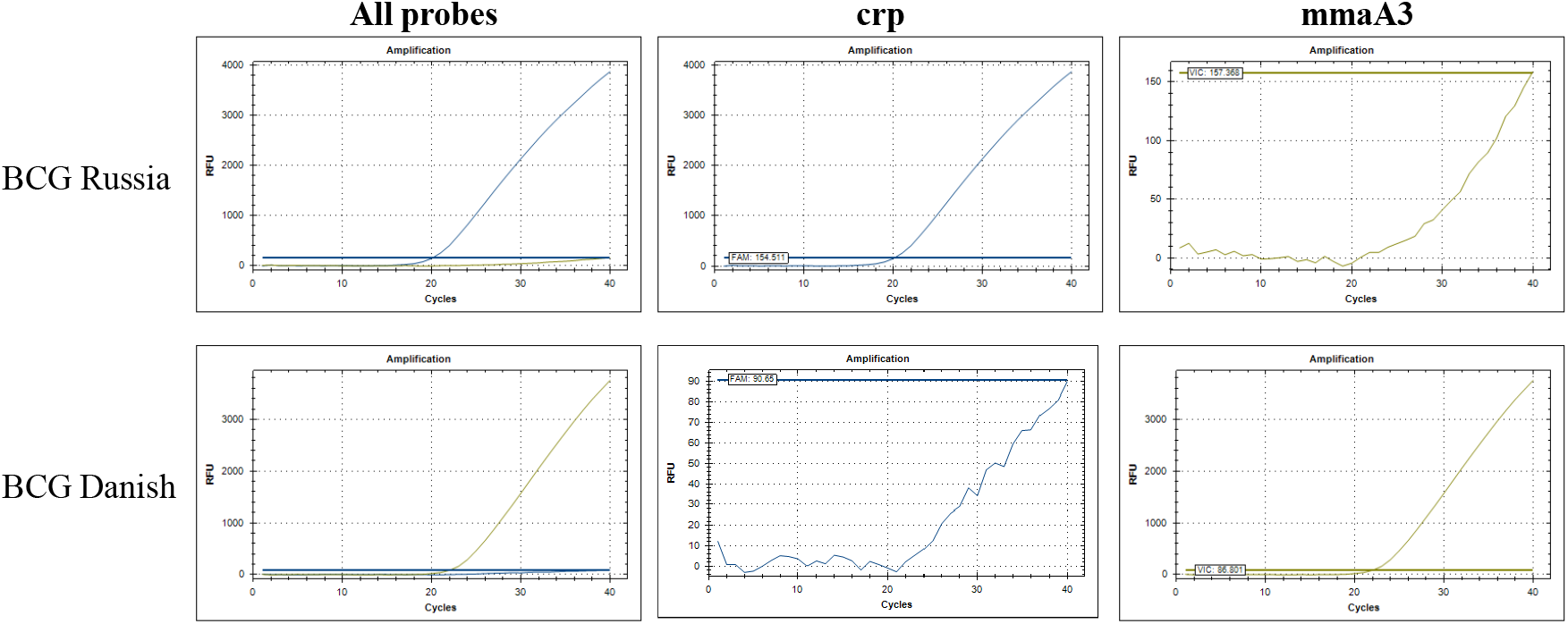
Step two assay specificity An example amplification curve for a BCG early strain BCG Russia and a late strain BCG Danish.

### Assay reproducibility and robustness

The inter- and intra-assay reproducibility was evaluated by comparing the Ct values of 3 separate plates with 5 technical replicates per plate. The Ct values remained consistent across all replicates: the Ct values remained under 2 Ct of difference across all replicates using step one (Fig. S4A) and under 3 Ct of difference across all replicates using step two (Fig. S4B). The ability of all probes to produce the expected amplification plot in the presence of excess non-specific DNA was tested using 1 ng target DNA (either BCG Russia or BCG Danish as indicated) with the addition of 10 ng of DNA from *M. abscessus*. All probes amplified as expected despite the presence of non-specific DNA (Fig. S5).

### Identification from mice

To determine whether the assay could be used to detect BCG directly from tissue, 3 C57BL/6 mice were intravenously infected with *M. bovis* BCG Russia. The mice were sacrificed 3 weeks post-infection and DNA was extracted from the spleen and liver. In all samples, the step one assay correctly identified the presence of BCG DNA (Fig. 3A) and the step two assay correctly identified the presence of an early BCG strain (Fig. 3B).

**Figure 3:**
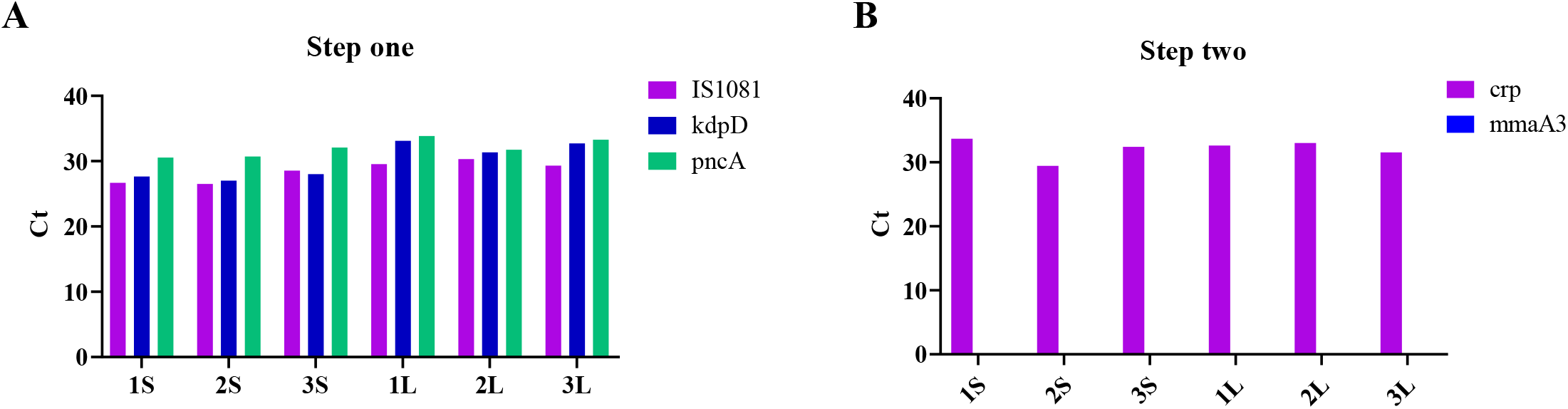
Real-time PCR results of DNA extracted directly from tissue of BCG-infected mice (A, B) The Ct values of DNA analyzed by the step one (A) and step two (B) assays. The number indicates the mouse, S indicates DNA extracted from spleen, and L indicates DNA extracted from liver.

### Validation with clinical samples

The assay was validated on a set of 19 isolates previously collected at Christian Medical College in Vellore, India (Table 2). Isolates were analyzed from patients with suspected tuberculosis who were 2 years of age or younger or who were suspected to have localised BCG infection or disseminated BCG due to a known immune condition. Samples were taken from pus (6), lymph node (4), gastric aspirate (4), thigh sinus tract (1), bone (1), pleural tissue (1), colonic ulcer (1), or mastoid tissue (1). Samples came from patients from 5 states in India: 8 from Tamil Nadu, 5 from Andhra Pradesh, 2 from Karnataka, 2 from West Bengal, and 1 from Odisha. One sample was taken from a patient from Bangladesh. DNA was extracted from culture on MGIT (13) or LJ medium (5), or directly from a pus swab (1) (Table S1). Of the 19 isolates included, 10 (52.6%) were confirmed to be BCG. Four (40%) of the BCG isolates were from female patients. The median age of BCG-infected patients was 6.5 months (range 3-12 months). In total, 4 of the BCG isolates came from patients with diagnosed with immune conditions: 2 BCG isolates were from patients with SCID and 2 from those with MSMD. Of the 10 BCG isolates, 6 (60%) were identified as a BCG early strain and 4 (40%) were identified as a BCG late strain. Example amplification plots of 2 clinical isolates, 1 early and 1 late strain, are found in figure S6. The other 9 isolates included in this study were identified as MTBC by the step one assay. Two (∼22.22%) of the MTBC isolates came from female patient samples. Of the 9 MTBC isolates, 7 (∼77.78%) came from patients between 1 and 2 years old.

**Table 2:**
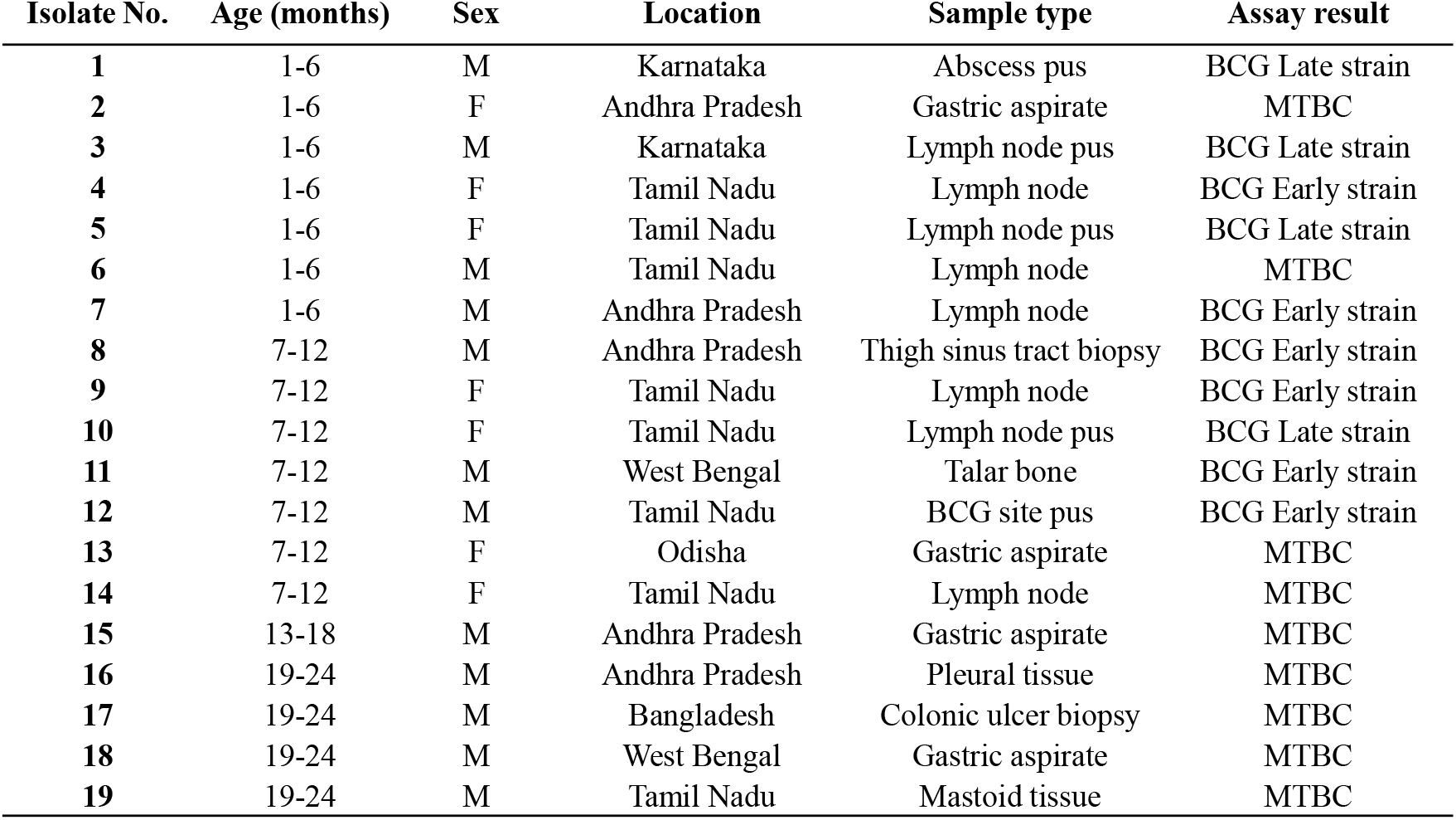
Patient information and assay results of 19 suspected BCG clinical isolates. Ages are presented as 6 month ranges.

| Isolate No. | Age (months) | Sex | Location | Sample type | Assay result |
| --- | --- | --- | --- | --- | --- |
| 1 | 1-6 | M | Karnataka | Abscess pus | BCG Late strain |
| 2 | 1-6 | F | Andhra Pradesh | Gastric aspirate | MTBC |
| 3 | 1-6 | M | Karnataka | Lymph node pus | BCG Late strain |
| 4 | 1-6 | F | Tamil Nadu | Lymph node | BCG Early strain |
| 5 | 1-6 | F | Tamil Nadu | Lymph node pus | BCG Late strain |
| 6 | 1-6 | M | Tamil Nadu | Lymph node | MTBC |
| 7 | 1-6 | M | Andhra Pradesh | Lymph node | BCG Early strain |
| 8 | 7-12 | M | Andhra Pradesh | Thigh sinus tract biopsy | BCG Early strain |
| 9 | 7-12 | F | Tamil Nadu | Lymph node | BCG Early strain |
| 10 | 7-12 | F | Tamil Nadu | Lymph node pus | BCG Late strain |
| 11 | 7-12 | M | West Bengal | Talar bone | BCG Early strain |
| 12 | 7-12 | M | Tamil Nadu | BCG site pus | BCG Early strain |
| 13 | 7-12 | F | Odisha | Gastric aspirate | MTBC |
| 14 | 7-12 | F | Tamil Nadu | Lymph node | MTBC |
| 15 | 13-18 | M | Andhra Pradesh | Gastric aspirate | MTBC |
| 16 | 19-24 | M | Andhra Pradesh | Pleural tissue | MTBC |
| 17 | 19-24 | M | Bangladesh | Colonic ulcer biopsy | MTBC |
| 18 | 19-24 | M | West Bengal | Gastric aspirate | MTBC |
| 19 | 19-24 | M | Tamil Nadu | Mastoid tissue | MTBC |

## Discussion

A rapid method of differentiating BCG from other mycobacterial species is essential to providing optimal care for BCG infection, particularly in cases of disseminated BCG which is associated with a mortality rate of ∼60%^4, 18-23^. A method of determining whether an early or late strain is causing a case of disseminated BCG may also provide an opportunity for pharmacosurveillance, to determine whether certain strains are linked to a higher rate of adverse events^2, 24^. Here, we have developed a two-step multiplex real-time PCR assay to rapidly identify BCG isolates from other mycobacterial species and differentiate between BCG early and late strains. This assay uses a rule-in approach to differentiate mycobacterial isolates on the basis of SNPs, to avoid potentially unreliable RD-based typing^11-14^. This assay was shown to detect BCG from culture, directly from tissue of infected mice, or from a patient pus swab in a case where culture was unavailable. The ability to identify BCG directly from a clinical sample circumvents time-consuming mycobacterial culture to inform patient treatment. The use of this assay was validated on 19 clinical isolates previously collected at Christian Medical College in Vellore, India, where two different BCG strains are in use: BCG Russia (an early strain) and BCG Danish (a late strain). Ten out of the 19 isolates were identified as BCG, 6 of which were early strains.

The clinical management of a patient diagnosed with BCG disease will differ compared to a patient with *M. tuberculosis*. Confirmation of BCG infection may affect which drugs would be included in the treatment regimen. Since BCG is derived from *M. bovis*, all strains are naturally resistant to pyrazinamide, a first-line anti-tuberculosis drug^16^. The assay described in this study confirms pyrazinamide resistance using the pncA probe. In addition, the mmaA3 probe used to identify late strains detects a SNP which is associated with a loss of methoxymycolate production and is suggested to cause low levels of isoniazid resistance in late strains^16, 26^. A diagnosis of BCG infection should also lead to further investigation of possible underlying immune conditions, which may not have been recognized at the time of disease presentation. In a previous study of 349 SCID patients, 118 developed disseminated BCG disease, a 33,000-fold increase over the general population^18^. In this study, of the 10 isolates identified as BCG, 2 came from patients confirmed to have SCID and 2 came patients confirmed to have MSMD. Although BCG vaccination is contraindicated in people with these conditions, the universal immunization programme (UIP) recommends BCG vaccination in India at birth or as early as possible (https://www.nhp.gov.in/universal-immunisation-programme_pg), so these rare immune conditions are typically not recognized until after vaccination.

There are 14 different BCG strains which have evolved with considerable genetic and phenotypic heterogeneity among them. BCG strains differ from one another by genetic deletions, SNPs and duplications^27^. It has been suggested that some BCG strains may be associated with a higher rate of adverse events^27^. This possibility is of particular interest in India where both an early and a late BCG strain are currently in use. In Finland, BCG osteitis rates were as high as 36.9 per 100,000 while using the early strain BCG Sweden. Upon switching to the late strain BCG Glaxo in 1978, that rate decreased to 6.4 per 100,000^2, 24^. Conversely, in phase one of a randomized control trial where BCG Danish was compared with BCG Russia, lymphadenitis was reported in 6 out of 582 patients given BCG Danish and one out of 575 patients given BCG Russia^28^. In this study, 6 out of 10 BCG isolates were early strains, presumably BCG Russia. The other 4 were identified as late strains, presumably BCG Danish. The sample size of this study is too small to draw conclusions concerning an association between BCG strain and adverse events, however screening is ongoing, and this question may be re-visited with the established assay as total sample size rises.

To conclude, we have developed a rapid method of reliably identifying BCG early and late strains which improves upon previously described assays. This assay may be used to inform patient treatment without the time constraints of mycobacterial culture. The ability to differentiate between early and late strains may also inform BCG vaccine pharmacovigilance in a country in which two vaccines strains are currently in use.

## Data Availability

All data referred to in the manuscript is available in the main text and supplementary material.

## Acknowledgements

This research was supported by the Canadian Institutes for Health Research (grant FDN148362 to M.A.B) and the Bill & Melinda Gates Foundation (OPP1176950 to V.K., M.A.B and J.S.M). The funding agency had no role in the study design, data collection, analysis, interpretation, or decision to submit this work for publication. We would like to thank Fiona McIntosh and Sarah Danchuk for technical support with assay development.

## Legends

Figure S1: Assay workflow and interpretation

Figure S2: Reaction efficiency calculations of step one probes

(A-C) A plot of the Ct values of a 10-fold serial dilution of BCG Russia DNA and calculation of the reaction efficiency for the IS1081 probe (A), the kdpD probe (B) and the pncA probe (C).

Figure S3: Reaction efficiency calculations of step two probes

(A) A plot of the Ct values of a 10-fold serial dilution of BCG Russia DNA and calculation of the reaction efficiency of the crp probe. (B) A plot of the Ct values of a 10-fold serial dilution of BCG Danish DNA and calculation of the reaction efficiency of the mmaA3 probe.

Figure S4: Inter- and Intra-assay reproducibility

(A, B) The Ct values were compared using 1ng of BCG Russia DNA with step 1 (A) or step 2 (B) assay. The inter-assay reproducibility was evaluated by comparing the Ct values across 5 technical replicates. The intra-assay reproducibility was evaluated by comparing the Ct values across 3 plates.

Figure S5: Assay performance in presence of 10ng of excess non-specific DNA

The ability of the assay to produce the expected amplification plot in the presence of excess non-specific DNA was evaluated. (A) A total of 10ng of DNA from *M. abscessus* (*Mab*) was added to a step one reaction mixture containing 1ng of BCG Russia DNA. (B) A total of 10ng of DNA from *Mab* was added to a step one reaction mixture containing 1ng of either BCG Russia or BCG Danish DNA.

Table S1: DNA extraction information for 19 suspected BCG clinical isolates MGIT – mycobacterial growth indicator tube, LJ – Lowenstein Jensen medium, MTBC – *Mycobacterium tuberculosis* complex

Figure S6: Clinical isolates amplification plots

(A, B) Example amplification plots of 2 clinical isolates using the step one (A) and step two (B) assay. Isolate number 3 was identified as a BCG late strain and isolate number 4 was identified as a BCG early strain.

