## Supplemental Fig 1-6, Supplemental Table 1 for "Development of a multiplex real-time PCR assay for BCG and validation in a clinical laboratory"

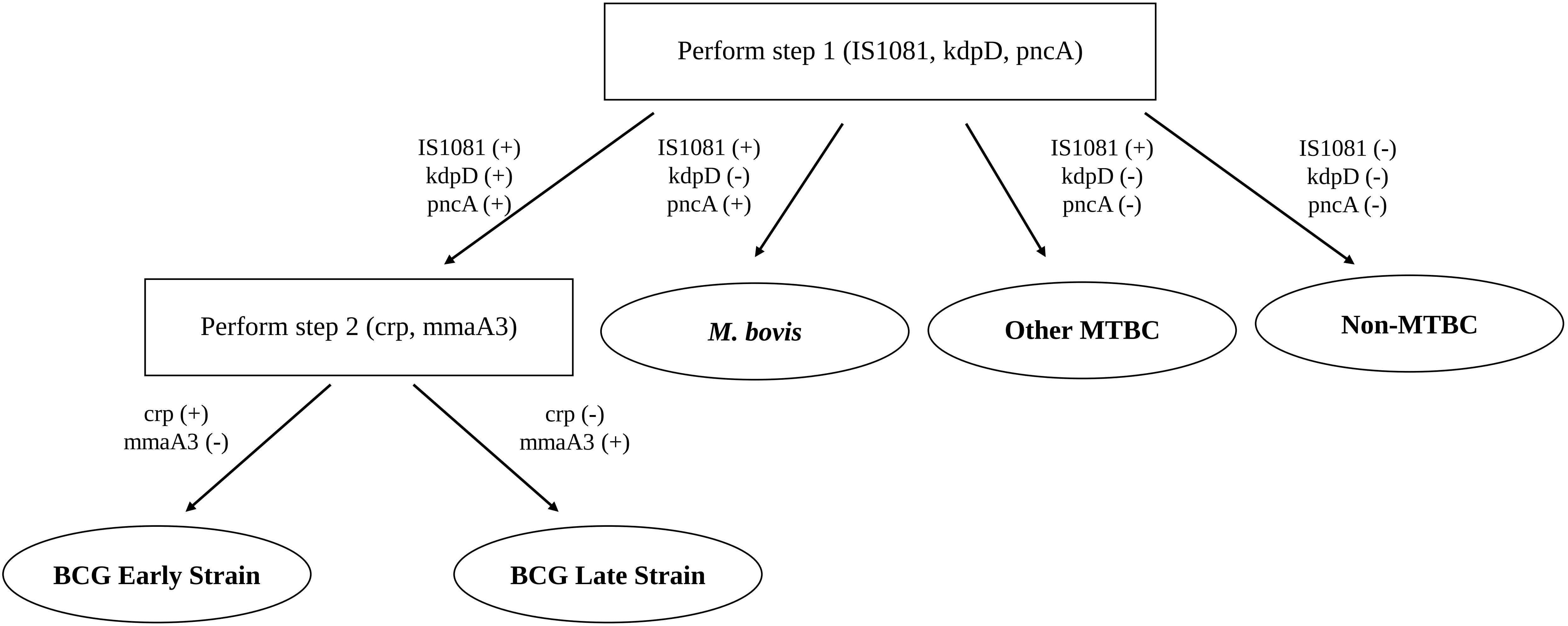


**Figure S1:** Assay workflow and interpretation

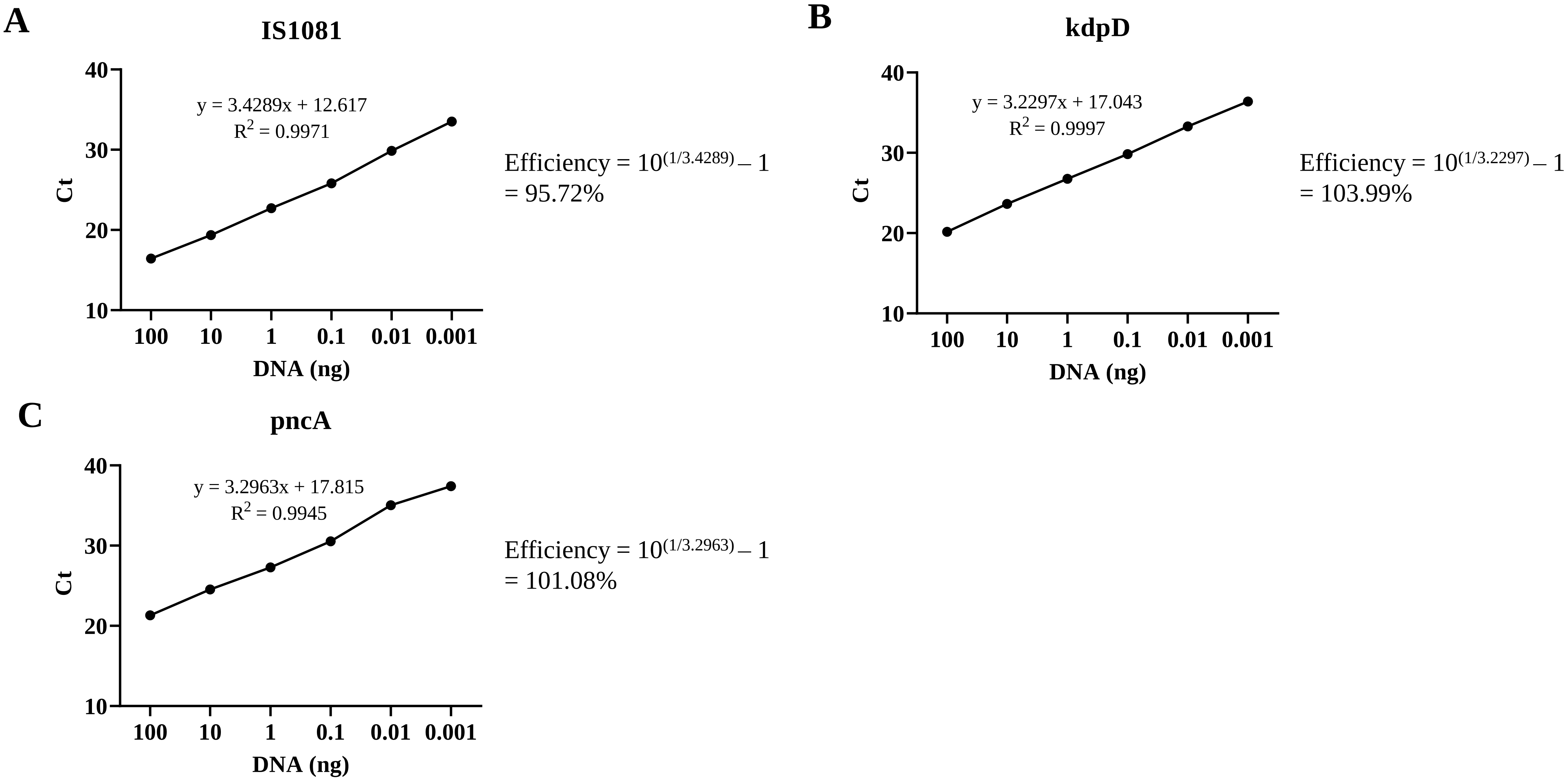


**Figure S2:** Reaction efficiency calculations of step one probes

(A-C) A plot of the Ct values of a 10-fold serial dilution of BCG Russia DNA and calculation of the reaction efficiency for the IS1081 probe (A), the kdpD probe (B) and the pncA probe (C).

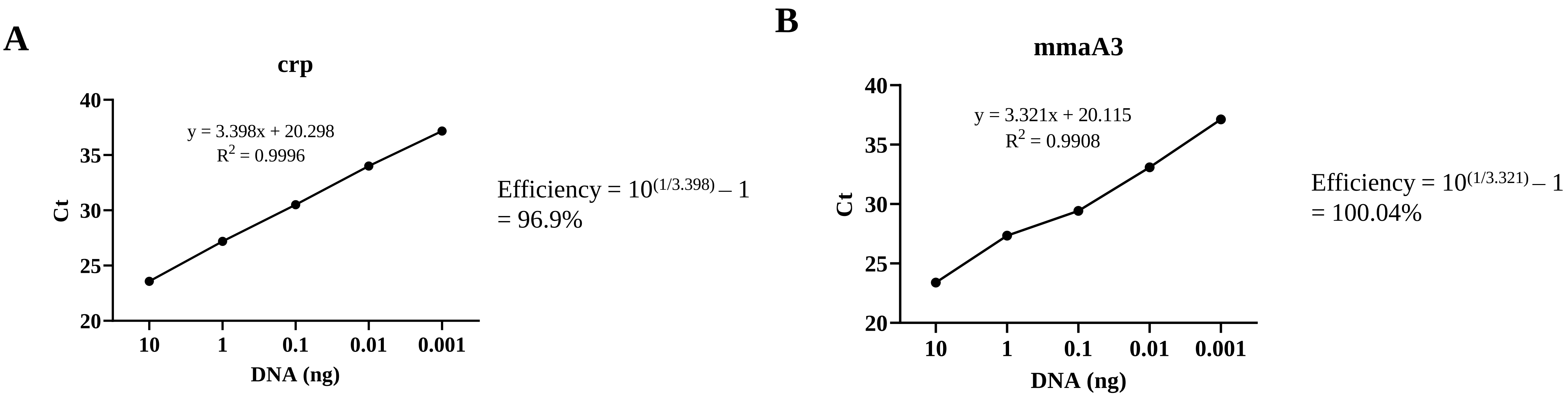


**Figure S3:** Reaction efficiency calculations of step two probes
(A) A plot of the Ct values of a 10-fold serial dilution of BCG Russia DNA and calculation of the reaction efficiency of the crp probe. (B) A plot of the Ct values of a 10-fold serial dilution of BCG Danish DNA and calculation of the reaction efficiency of the mmaA3 probe.

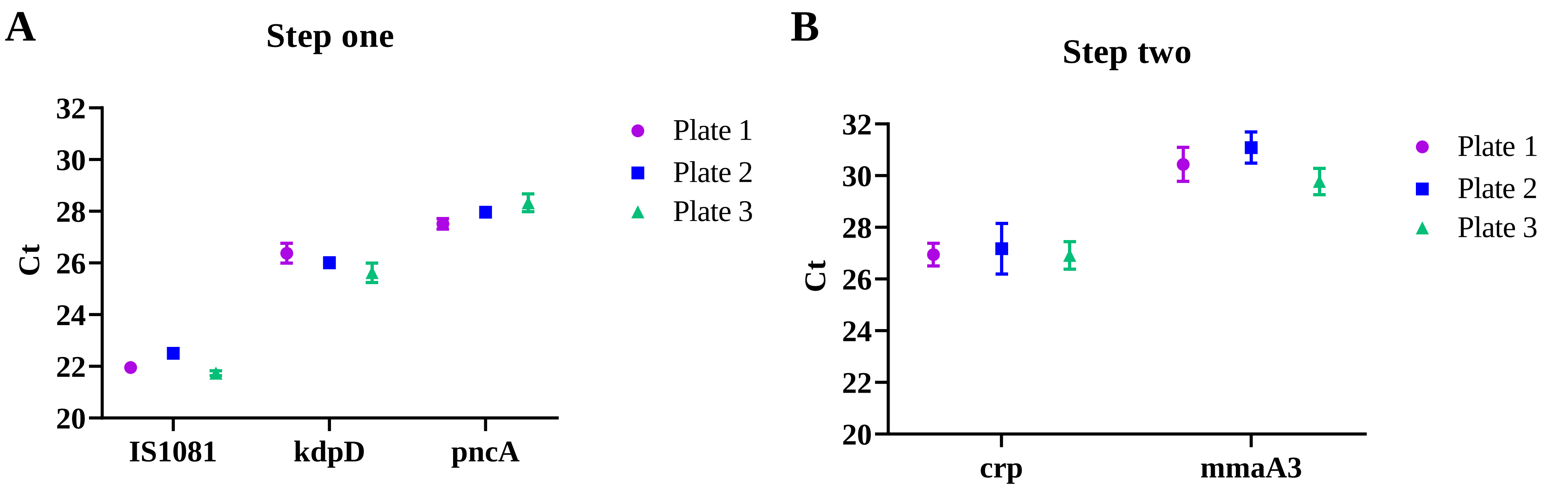


**Figure S4**: Inter- and Intra-assay reproducibility
(A, B) The Ct values were compared using 1ng of BCG Russia DNA with step 1 (A) or step 2 (B) assay. The inter-assay reproducibility was evaluated by comparing the Ct values across five technical replicates. The intra-assay reproducibility was evaluated by comparing the Ct values across three assay replicates.

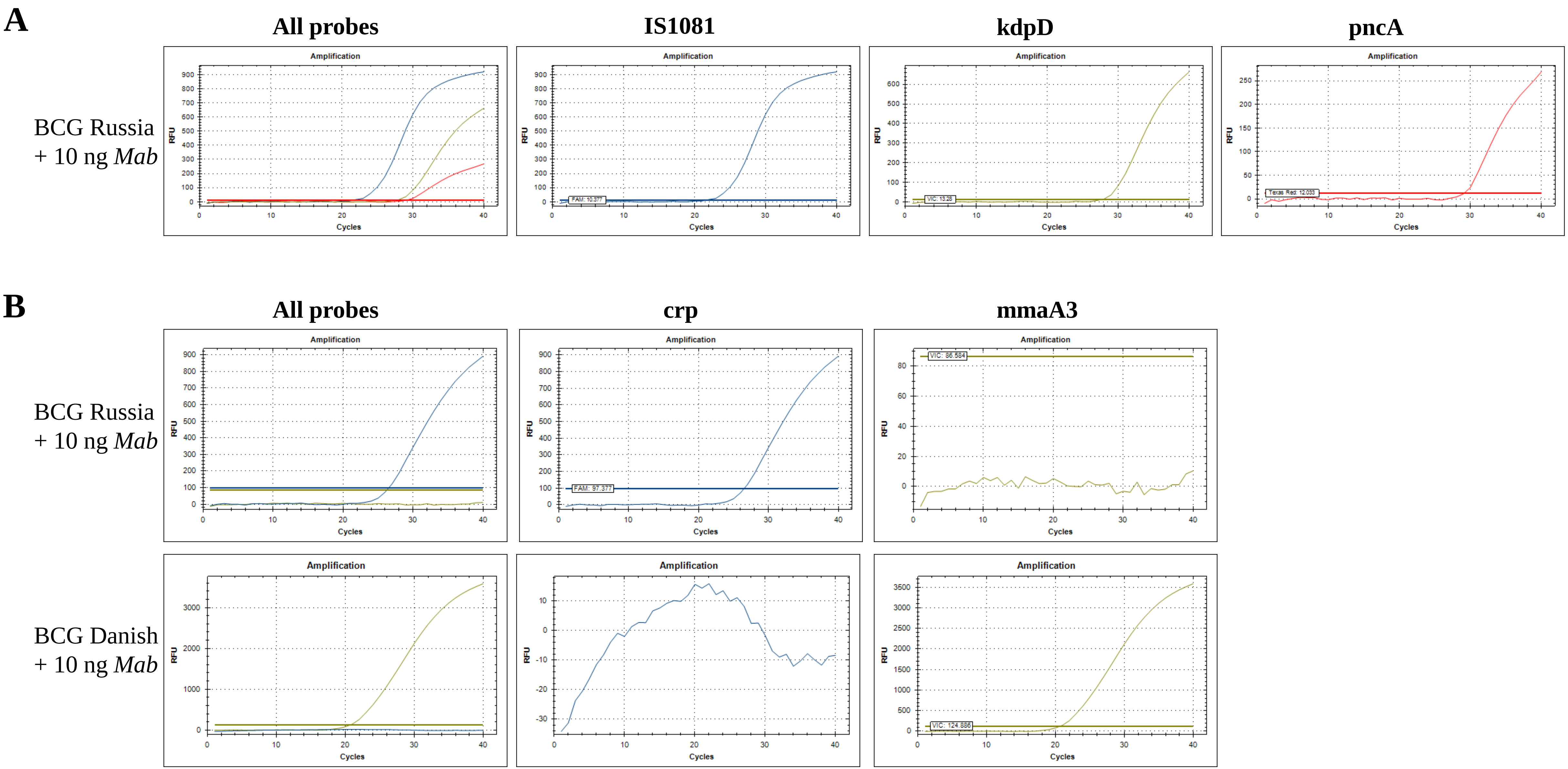


**Figure S5**: Assay performance in presence of 10ng of excess non-specific DNA

The ability of the assay to produce the expected amplification plot in the presence of excess non-specific DNA was evaluated. (A) A total of 10ng of DNA from *M. abscessus* (*Mab*) was added to a step one reaction mixture containing 1ng of BCG Russia DNA. (B**)** A total of 10ng of DNA from *Mab* was added to a step one reaction mixture containing 1ng of either BCG Russia or BCG Danish DNA.

| Isolate No. | Age (months) | Sex | Location | Sample type | Year of diagnosis | Extraction from | Extraction method | Assay result |
|---|---|---|---|---|---|---|---|---|
| 1 | 1-6 | M | Karnataka | Abscess pus | 2020 | LJ | Qiagen kit | BCG Late strain |
| 2 | 1-6 | F | Andhra Pradesh | Gastric aspirate | 2020 | MGIT | Qiagen kit | MTBC |
| 3 | 1-6 | M | Karnataka | Lymph node pus | 2019 | LJ | Qiagen kit | BCG Late strain |
| 4 | 1-6 | F | Tamil Nadu | Lymph node | 2018 | MGIT | Heat prep | BCG Early strain |
| 5 | 1-6 | F | Tamil Nadu | Lymph node pus | 2020 | LJ | Qiagen kit | BCG Late strain |
| 6 | 1-6 | M | Tamil Nadu | Lymph node | 2019 | MGIT | Heat prep | MTBC |
| 7 | 1-6 | M | Andhra Pradesh | Lymph node | 2018 | MGIT | Heat prep | BCG Early strain |
| 8 | 7-12 | M | Andhra Pradesh | Thigh sinus tract biopsy | 2020 | LJ | Qiagen kit | BCG Early strain |
| 9 | 7-12 | F | Tamil Nadu | Lymph node pus | 2019 | MGIT | Heat prep | BCG Early strain |
| 10 | 7-12 | F | Tamil Nadu | Lymph node pus | 2020 | Pus swab | Qiagen kit | BCG Late strain |
| 11 | 7-12 | M | West Bengal | Talar bone | 2019 | MGIT | Qiagen kit | BCG Early strain |
| 12 | 7-12 | M | Tamil Nadu | BCG site pus | 2018 | MGIT | Heat prep | BCG Early strain |
| 13 | 7-12 | F | Odisha | Gastric aspirate | 2020 | MGIT | Qiagen kit | MTBC |
| 14 | 7-12 | F | Tamil Nadu | Lymph node | 2020 | MGIT | Qiagen kit | MTBC |
| 15 | 13-18 | M | Andhra Pradesh | Gastric aspirate | 2019 | MGIT | Heat prep | MTBC |
| 16 | 19-24 | M | Andra Pradesh | Pleural tissue | 2019 | MGIT | Heat prep | MTBC |
| 17 | 19-24 | M | Bangladesh | Colonic ulcer biopsy | 2019 | MGIT | Heat prep | MTBC |
| 18 | 19-24 | M | West Bengal | Gastric aspirate | 2019 | MGIT | Qiagen kit | MTBC |
| 19 | 19-24 | M | Tamil Nadu | Mastoid tissue | 2020 | LJ | Qiagen kit | MTBC |

**Table S1:** DNA extraction information for 19 suspected BCG clinical isolates
MGIT – mycobacterial growth indicator tube, LJ – Lowenstein Jensen medium, MTBC – *Mycobacterium tuberculosis* complex

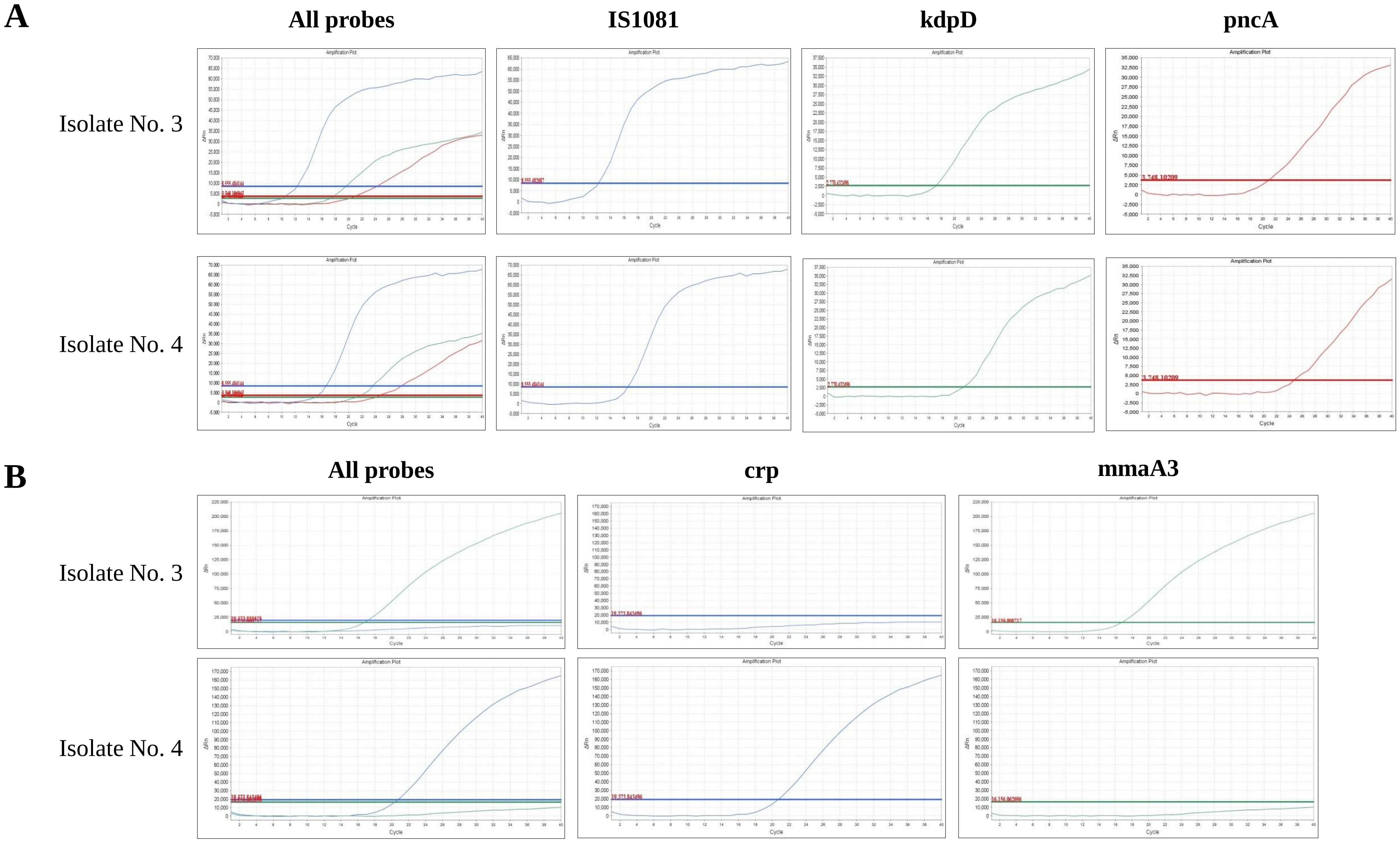


**Figure S6**: Clinical isolates amplification plots

(A, B) Example amplification plots of two clinical isolates using the step one (A) and step two (B) assay. Isolate number 3 was identified as a BCG late strain and isolate number 4 was identified as a BCG early strain.
